# Evaluation of a novel community-based COVID-19 ‘Test-to-Care’ model for low-income populations

**DOI:** 10.1101/2020.07.28.20161646

**Authors:** Andrew D. Kerkhoff, Darpun Sachdev, Sara Mizany, Susy Rojas, Monica Gandhi, James Peng, Douglas Black, Diane Jones, Susana Rojas, Jon Jacobo, Valerie Tulier-Laiwa, Maya Petersen, Jackie Martinez, Gabriel Chamie, Diane V. Havlir, Carina Marquez

**Affiliations:** Division of HIV, Infectious Diseases, and Global Medicine, Department of Medicine, University of California, San Francisco, San Francisco, CA, USA; San Francisco Department of Public Health, San Francisco, CA, USA; The San Francisco Latino Task Force on COVID-19; Division of Epidemiology and Biostatistics, the School of Public Health, University of California, Berkeley, Berkeley, CA, USA

**Keywords:** COVID-19, community health workers, promotores, social support, community-based, vulnerable populations, Latinx, RE-AIM

## Abstract

**Background:** After a COVID-19 diagnosis, vulnerable populations face considerable logistical and financial challenges to isolate and quarantine. We developed and evaluated a novel, community-based approach (‘Test-to-Care’ Model) designed to address these barriers for socioeconomically vulnerable Latinx individuals with newly diagnosed COVID-19 and their households.

**Methods:** This three-week demonstration project was nested within an epidemiologic surveillance study in a primarily Latinx neighborhood in the Mission district of San Francisco, California. The Test-to-Care model was developed with input from community members and public health leaders. Key components included: (1) provision of COVID-19-related education and information about available community resources, (2) home deliveries of material goods to facilitate safe isolation and quarantine (groceries, personal protective equipment and cleaning supplies), and (3) longitudinal clinical and social support. Newly SARS-CoV-2 PCR-positive participants were eligible to participate. Components of the model were delivered by the Test-to-Care team which was comprised of healthcare providers and community health workers (CHWs) who provided longitudinal clinic- and community-based support for the duration of the isolation period to augment existing services from the Department of Public Health (DPH). We evaluated the Test-to-Care Model using the Reach, Effectiveness, Adoption, Implementation, Maintenance (RE-AIM) Framework and drew upon multiple data sources including: programmatic data, informal interviews with participants and providers/CHWs and structured surveys among providers/CHWs.

**Results:** Overall, 83 participants in the surveillance study were diagnosed with COVID-19, of whom 95% (79/83) were Latinx and 88% (65/74) had an annual household income <$50,000. Ninety-six percent (80/83) of participants were reached for results disclosure, needs assessment and DPH linkage for contact tracing. Among those who underwent an initial needs assessment, 45% (36/80) were uninsured and 55% (44/80) were not connected to primary care. Sixty-seven percent (56/83) of participants requested community-based CHW support to safely isolate at their current address and 65% (54/83) of all COVID-19 participants received ongoing community support via CHWs for the entire self-isolation period. Participants reported that the intervention was highly acceptable and that their trust increased over time – this resulted in 9 individuals who disclosed a larger number of household members than first reported, and 6 persons who requested temporary relocation to a hotel room for isolation despite initially declining this service; no unintended harms were identified. The Test-to-Care Model was found to be both acceptable and feasible to providers and CHWs. Challenges identified included a low proportion of participants linked to primary care despite support (approximately 10% after one month), and insufficient access to financial support for wage replacement.

**Conclusions:** The Test-to-Care Model is a feasible and acceptable intervention for supporting self-isolation and quarantine among newly diagnosed COVID-19 patients and their households by directly addressing key barriers faced by vulnerable populations.

## Introduction

COVID-19 disproportionately affects racial and ethnic minorities in the United States [1–4] In San Francisco, Latinx individuals comprise 50% of the COVID-19 cases, despite making up 15% of the population [5]. Pre-existing structural inequities drive this excess number of cases in the Latinx community [4,6]. Following diagnosis with COVID-19, low-income Latinx and other socioeconomically vulnerable populations face considerable logistical and financial challenges to safely isolate and quarantine. Without the availability of a comprehensive model of care that can overcome barriers to the required isolation and quarantine, including: lack of access to culturally-tailored COVID-19 education, lack of access to food and personal protective equipment, and lack of social support, as well as the potentially catastrophic financial consequences, low-income persons are unlikely to undertake testing for COVID-19. Furthermore, due to fear and a lack of trust, individuals may be unwilling to name household and other close contacts, especially if they are undocumented. To address known health disparities and to ensure that test, isolate and trace strategies for COVID-19 are reaching the most vulnerable and affected persons, there is an urgent need to develop and evaluate tailored low-barrier testing strategies paired with social support interventions during the isolation period [7,8].

Community health workers (CHWs), also known as promotores de salud, have been increasingly utilized in medical and public health interventions to engage marginalized and traditionally hard-to-reach individuals and reduce health disparities [9–12]. They often share the same language, ethnicity, community and/or life experiences as the individuals they serve; thus, their involvement in the design and implementation of health interventions can help to ensure cultural relevance through alignment with local concerns and priorities. CHWs can overcome individuals’ barriers to engagement and retention in health services through several mechanisms including by serving as a trusted and credible source for healthcare information, increasing social support, reducing stigma, and through sharing of health-specific knowledge. Several studies have previously demonstrated that CHW interventions improved health behaviors and outcomes across a number of health domains, including diabetes [13,14], hypertension [15], asthma [16], cancer prevention [17] mental health [18] and HIV [19,20]; CHWs have also shown to be both effective and cost-effective for improving health outcomes specifically among underserved individuals and racial and ethnic minority communities [17,21]. Despite urgent calls to scale-up CHWs to help respond to the COVID-19 pandemic [22–25], to date there have been no published evaluations of interventions that have incorporated community-based support provided by CHWs into a comprehensive care model to respond to and support the complex needs of low-income individuals affected by COVID-19.

We have previously reported on the results of an epidemiologic surveillance study (Unidos en Salud) conducted from April 25-28, 2020 in the Mission Neighborhood of San Francisco California that found that the point prevalence of PCR-positive SARS-CoV-2 infection was 20-times higher among Latinx residents compared to non-Latinx residents (3.9% vs. 0.2%) [26]. Notably, 96% of recent COVID-19 infections were among Latinx individuals who were predominantly low-income, lived in densely populated households and were frontline workers or unemployed persons who could not afford to shelter-in-place. To overcome the substantial barriers to self-isolation and quarantine faced by socioeconomically vulnerable individuals and their household members, we developed a model of enhanced clinic- and community-based support, including longitudinal support from CHWs (Test-to-Care Model [T2C]) for individuals who tested SARS-CoV-2 PCR-positive during the Unidos en Salud study. In this paper we describe the development of the T2C Model and evaluate its reach, feasibility and acceptability.

## Methods

### Setting

The community-based COVID-19 testing campaigns and subsequent test-to-care demonstration project were undertaken in a single, densely populated, highly diverse, U.S. census tract (022901) within the Mission District of San Francisco, California. This represents a 16-square block area with approximately 5,174 residents of whom 58% are Latinx, 34% White/Caucasian, 5% Asian/Pacific Islander, and 1% Black/African American [26]. Many residents live in high density, low-income households; the combined income of 34% of households in the district is less than $50,000 per year. The Mission District is historically a predominantly Latinx district in San Francisco; however, over the last two decades it has undergone substantial gentrification, resulting in rapidly escalating housing costs and displacement of Latinx residents [27].

### Ethics

The UCSF Committee on Human Research determined that the study and subsequent program evaluation met criteria for public health surveillance, program evaluation and quality improvement activities, rendering it exempt from IRB oversight.

### Community-partnership approach

A community-academic partnership between the Latino Task Force for COVID-19 (LTF) and UCSF underpinned the design of Unidos en Salud (UeS) Study and all subsequent study activities, including the T2C model [28]. The Latino Task Force for COVID-19 consists of leaders from several long-standing Latinx community-based organizations that was formed to support and address the specific needs of the Latinx community in San Francisco during the COVID-19 pandemic [29]. Members of the Latino Task Force and the UCSF study team met several times a week to discuss successes, ongoing challenges and to engage in shared-decision making.

### Overview of the Test-to-Care Model

In conjunction with our community partners, we designed the T2C model to provide enhanced, longitudinal clinical and community-based support alongside the San Francisco Department of Public Health’s (SFDPH) case and contact investigation services to any participant of the Unidos en Salud Study who tested PCR-positive for SARS-CoV-2 (**Fig 1**) [26]. In addition to using a community partnership approach, the T2C model also drew upon our prior experiences and lessons learned from designing and implementing large scale programs to facilitate low-barrier HIV testing and linkage-to-care [30–32]The objectives of the T2C model were to: (1) provide longitudinal medical, social and emotional support to low-income individuals who tested positive for COVID-19 in order to address potential barriers for them and their households to safely self-isolate and quarantine throughout the duration of the recommended period of 10 days or more (**Fig 1, Table 1**); (2) provide direct and ongoing support to COVID-19 positive participants to enroll in health insurance and to establish or re-connect with a primary care provider and access community resources in order to create a foundation to support their positive physical and mental health beyond the demonstration project (**Fig 1, Table 1)**; and, (3) create an effective and sustainable community-based model utilizing CHWs that could be implemented in other communities to support the needs of low-income persons testing positive for COVID-19.

**Table 1.**
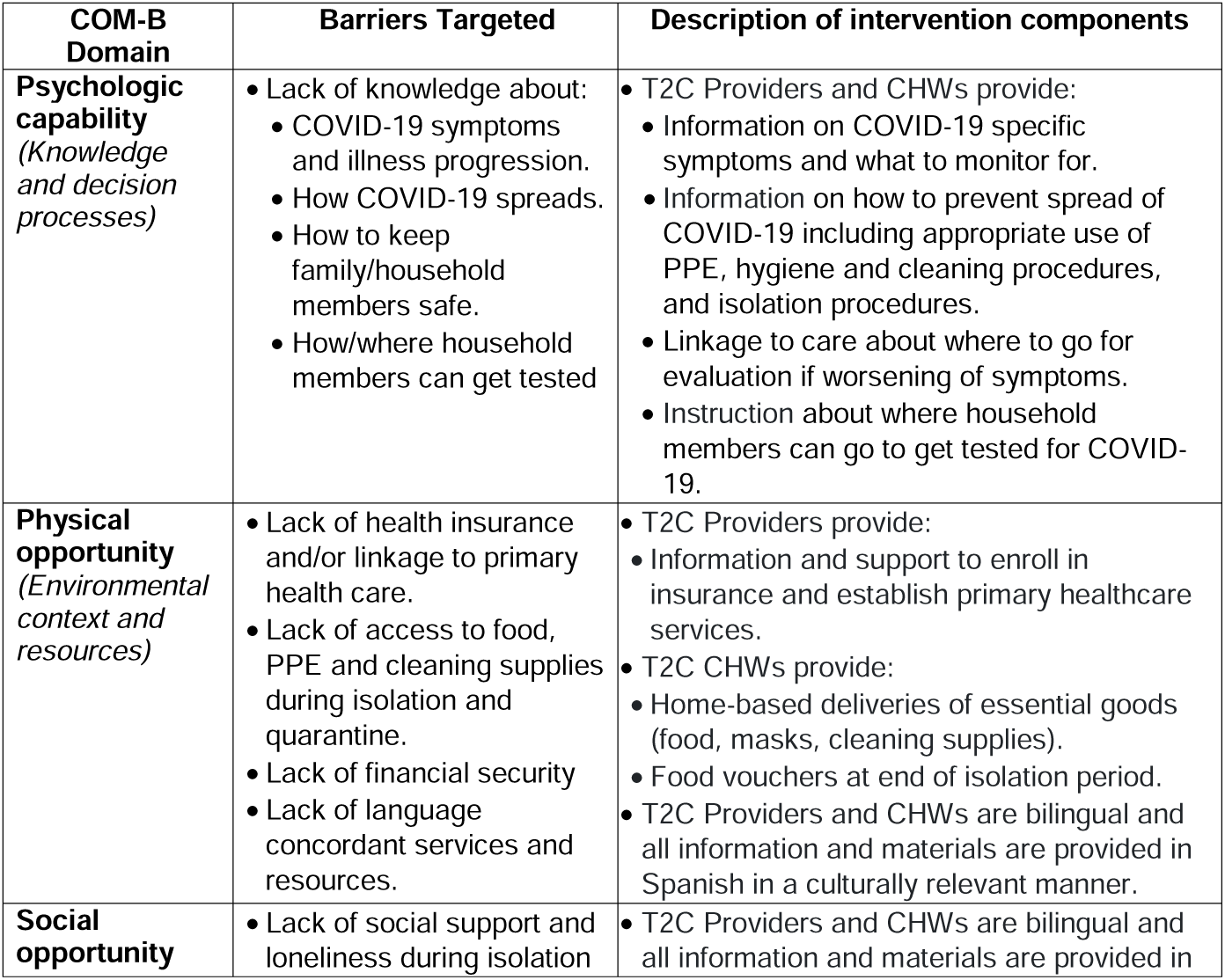
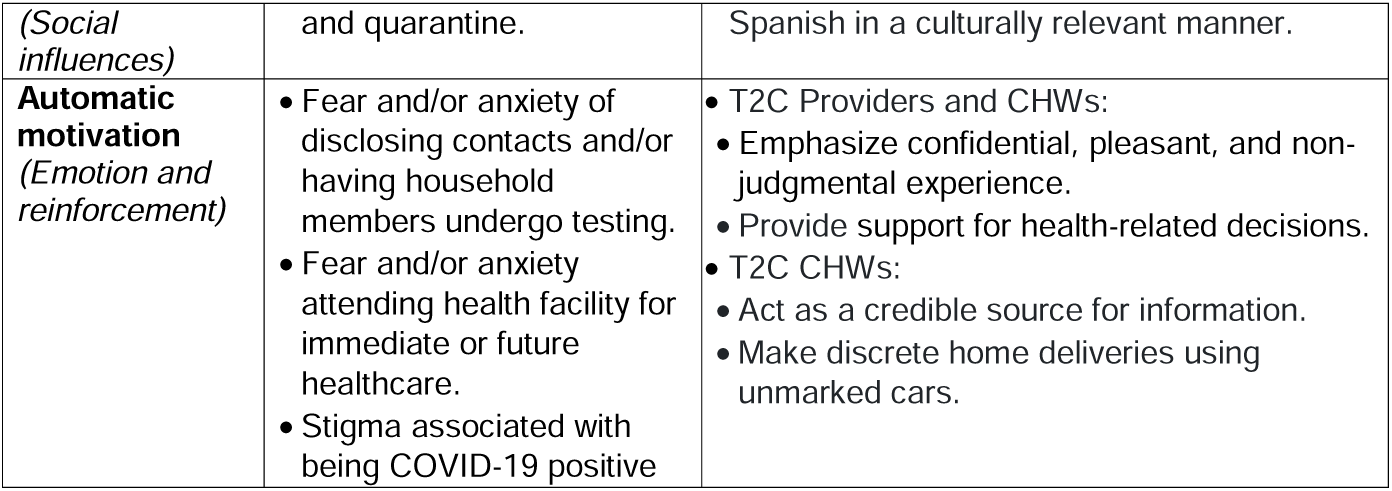
Hypothesized barriers faced by low-income, Latinx COVID-19 positive individuals and description of intervention components to address potential barriers. Barriers are categorized according to the Capability, Opportunity, and Motivation Model, which is a validated behavior change framework [33,34].

**Fig 1.**
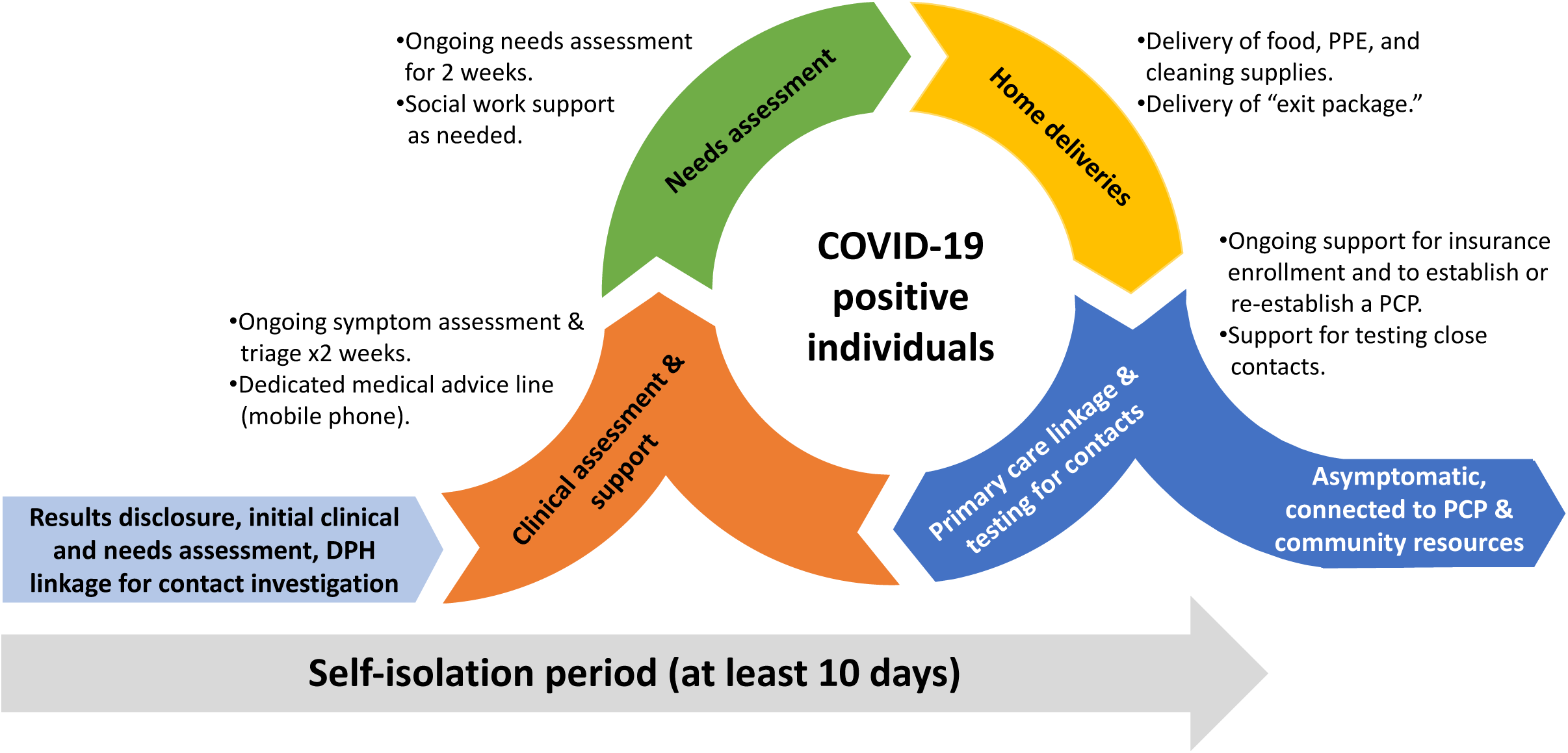
Overview of the Test-to-Care Model

### Description of the Test-to-Care Model Demonstration Project

We undertook a three-week demonstration project of the T2C Model from April 27^th^ to May 14^th^ that nested within the Unidos en Salud COVID-19 “Test and Respond” study [26]. The T2C model was carried out by a bilingual, multidisciplinary team comprised of CHWs and healthcare providers. There were three full-time, bilingual and bicultural community members who served as CHWs; these individuals had made extensive contributions as volunteers to prior Unidos en Salud study activities [26]. Healthcare providers included bilingual social workers, nurses and physicians who had extensive experience in providing care and support to highly vulnerable populations [31,32]. The T2C team had a community hub in the Mission neighborhood and a clinical hub at an outpatient clinic at San Francisco General Hospital.

The T2C huddled daily to discuss needs for initial and longitudinal support to COVID-19 positive individuals and their household members throughout the recommended period of self-isolation and quarantine, respectively **(Fig 1)**. Any participants testing COVID-19 PCR-positive were contacted by a bilingual clinician on the T2C team to provide same-day disclosure and to perform an initial screen to evaluate: (a) current symptoms and underlying medical comorbidities, (b) health insurance status, (c) primary care status, (d) ability to safely isolate at present address, (e) food security, (f) availability of and access to personal protective equipment (PPE) and cleaning supplies, and (g) other overt needs (**S1 Appendix**). Based upon this assessment, patients were categorized into one of three mutually exclusive categories according to the degree of support needed (low, medium and high), which determined the frequency of follow-up wellness calls made by the clinic-based T2C team (low: every 4-7 days, medium: every 3-4 days, high: every 1-2 days) (**S1 Appendix**).

During the initial disclosure phone call, participants were provided with information and education related to COVID-19 and how to safely self-isolate, as well as a dedicated phone number they could call during business hours if they had any questions or concerns. Participants were informed about the CHW-led, community-based T2C team and asked if they would like to be contacted and supported by this team. Individuals who agreed and needed food, personal protective equipment (PPE), and/or cleaning supplies to safely isolate and/or who wanted social support were contacted the same day by a CHW and were provided urgent home-delivery of essential goods (**Table 2**). A COVID-19 positive result and disclosure note was documented in the electronic medical record system and primary care providers were alerted of the result (if or when established). Work excuse notes were provided to participants when requested. The SFPDH undertook contact investigation in accordance with local procedures and practices; any COVID-19 PCR-positive participants unable to safely self-isolate at their current address were referred to isolation and quarantine hotel rooms established by the SFDPH. Isolation and quarantine hotel rooms were provided free of charge for the duration of the isolation period and were located outside of the Mission District, but within San Francisco.

**Table 2.**
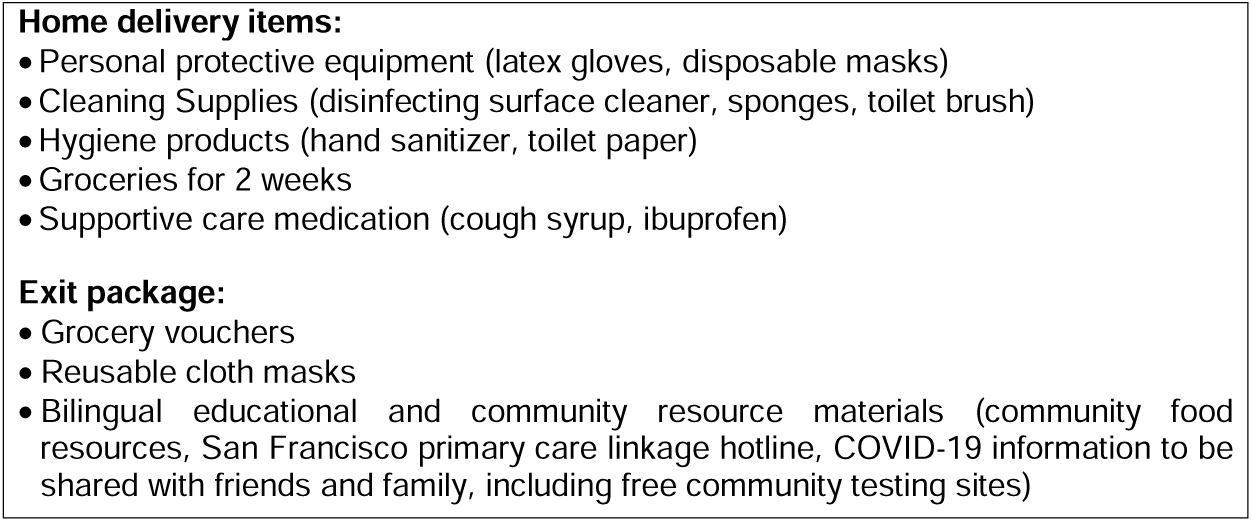
Overview of goods and products provided through home delivery by CHWs

For the remaining period of isolation, COVID-19 positive participants were followed by T2C team members to assess for new or unresolved needs and to provide ongoing support. Participants with new or worsening symptoms were triaged by a T2C provider and directed to urgent care or the emergency department as appropriate. Those without health insurance, without established primary care, or with food or financial insecurity were contacted by a T2C social worker to assess eligibility and provided information and ongoing support to link to appropriate community resources, including scheduling new patient appointments at community health clinics. CHWs continued to assess for food insecurity and the need for additional PPE and cleaning supplies among participants, which were addressed through regular home-deliveries (**Table 2**). CHWs also spent substantial time regularly talking with participants and their family members during the isolation and quarantine period, providing them with social support that consisted of: (a) continued education, advice and guidance, (b) emotional support, and (c) companionship, all of which served to develop trust among participants. To optimally support the needs of all participants, there was regular and frequent communication between all T2C members as well as the SFPDH when applicable. At the end of the self-isolation period all participants, independent of initial needs classification, were called to ensure that they were asymptomatic, or clinically improving. Additionally, participants who had been supported by CHWs were provided an “exit package” (**Table 2**).

### Evaluation of the Test-to-Care Model Demonstration Project

The Reach, Effectiveness, Adoption, Implementation and Maintenance (RE-AIM) framework was utilized to guide the evaluation of the T2C model demonstration project [27]. Reach was defined according to the number of COVID-19 positive participants successfully contacted for disclosure and initial needs assessment as well as the number of participants who wanted and were provided social work support and community-based support from CHWs. Preliminary effectiveness was evaluated according to several indicators, including: (1) participant self-report that the T2C model made it easier for them to safely self-isolate for the duration of the recommended period; (2) the number of household contacts identified, which may reflect participant trust of CHW; (3) the proportion of participants initially without health insurance and/or a primary care provider who established health insurance and a primary care provider after 4 weeks follow-up; (4) the proportion of participants with worsening symptoms who were triaged to appropriate evaluation/care; (5) the proportion of participants reached by CHWs who were followed and provided support for the entire period of self-isolation; and (6) any unintended consequences associated with the T2C model. Implementation measures included fidelity to the T2C model as intended, the perceived feasibility of the T2C model among T2C providers and CHWs, and acceptability of the T2C model among COVID-19 positive participants and the T2C team members.

A number of data sources informed the evaluation of the T2C model, including a brief structured survey among participants conducted on the initial date of COVID-19 testing [6], programmatic data from the T2C team, and data from the electronic medical record. The acceptability of the T2C model among participants was informed by informal interviews. Feasibility and acceptability of the T2C model among providers and CHWs was informed by both informal interviews as well as brief, structured surveys using five-point Likert scale questions (**S1 Table**) [35–37]. Participants were characterized using simple descriptive statistics. To further evaluate reach, participant characteristics of those wanting and receiving support from CHWs were compared to those who declined support from CHWs. The number of participants reached and supported by the T2C team was visually represented using a cascade-of-care analysis.

## Results

### Overview of COVID-19 positive participants

Among 3,871 residents and workers tested for COVID-19 from April 25-28, 2020, 83 were PCR-positive (prevalence, 2.1% [95%CI, 1.7-2.7]). We have previously reported the demographics of PCR-positive participants [26], but in brief, positive participants had a median age of 39 years, were predominantly male (76%) and nearly all were Latinx (95%) **(Table 3**). Two-thirds were frontline workers and 91% reported that they were unable to shelter-in-place and maintain their income. 81 (98%) were housed, while 2 (2%) were unhoused. Housed participants lived in one of 47 unique households with a median of 6 (IQR, 3-7) reported members per household; 88% of participants lived in a household with an annual income <$50,000 (**Table 3**).

**Table 3.**
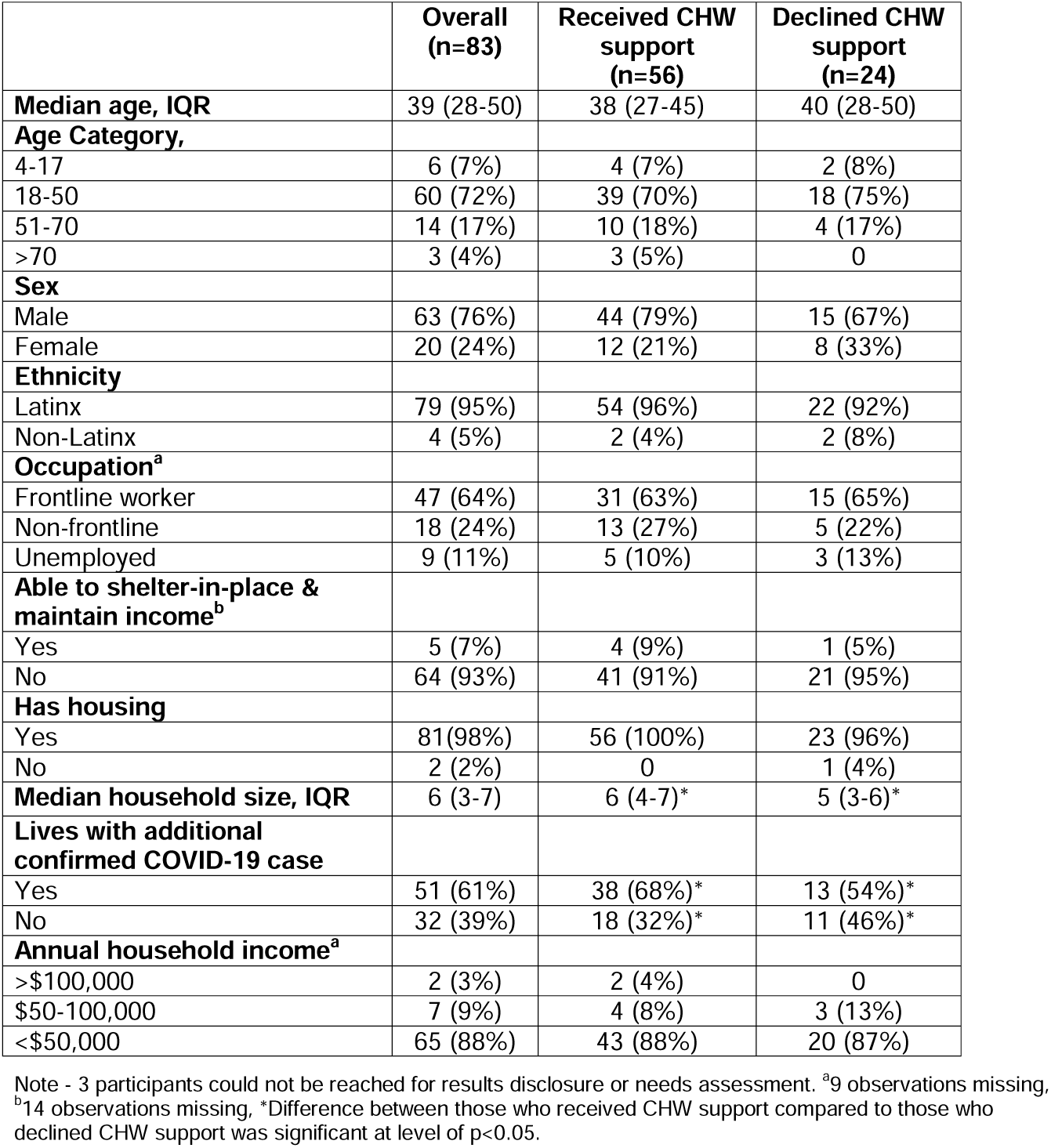
Baseline demographics and socioeconomic characteristics and reach of the CHW support component of the T2C model.

|  | **Overall**<br>**(n=83)** | **Received CHW**<br>**support**<br>**(n=56)** | **Declined CHW**<br>**support**<br>**(n=24)** |
| --- | --- | --- | --- |
| **Median age, IQR** | 39 (28-50) | 38 (27-45) | 40 (28-50) |
| **Age Category,** |  |  |  |
| 4-17 | 6 (7%) | 4 (7%) | 2 (8%) |
| 18-50 | 60 (72%) | 39 (70%) | 18 (75%) |
| 51-70 | 14 (17%) | 10 (18%) | 4 (17%) |
| >70 | 3 (4%) | 3 (5%) | 0 |
| **Sex** |  |  |  |
| Male | 63 (76%) | 44 (79%) | 15 (67%) |
| Female | 20 (24%) | 12 (21%) | 8 (33%) |
| **Ethnicity** |  |  |  |
| Latinx | 79 (95%) | 54 (96%) | 22 (92%) |
| Non-Latinx | 4 (5%) | 2 (4%) | 2 (8%) |
| **Occupation<sup>a</sup>** |  |  |  |
| Frontline worker | 47 (64%) | 31 (63%) | 15 (65%) |
| Non-frontline | 18 (24%) | 13 (27%) | 5 (22%) |
| Unemployed | 9 (11%) | 5 (10%) | 3 (13%) |
| **Able to shelter-in-place &**<br>**maintain income<sup>b</sup>** |  |  |  |
| Yes | 5 (7%) | 4 (9%) | 1 (5%) |
| No | 64 (93%) | 41 (91%) | 21 (95%) |
| **Has housing** |  |  |  |
| Yes | 81(98%) | 56 (100%) | 23 (96%) |
| No | 2 (2%) | 0 | 1 (4%) |
| **Median household size, IQR** | 6 (3-7) | 6 (4-7)* | 5 (3-6)* |
| **Lives with additional**<br>**confirmed COVID-19 case** |  |  |  |
| Yes | 51 (61%) | 38 (68%)* | 13 (54%)* |
| No | 32 (39%) | 18 (32%)* | 11 (46%)* |
| **Annual household income<sup>a</sup>** |  |  |  |
| >\$100,000 | 2 (3%) | 2 (4%) | 0 |
| \$50-100,000 | 7 (9%) | 4 (8%) | 3 (13%) |
| <\$50,000 | 65 (88%) | 43 (88%) | 20 (87%) |

Note - 3 participants could not be reached for results disclosure or needs assessment. <sup>a</sup>9 observations missing, <sup>b</sup>14 observations missing, \*Difference between those who received CHW support compared to those who declined CHW support was significant at level of p<0.05.

## Reach

### Result of disclosure and needs assessment

Of 83 COVID-19 positive participants, 80 (96%) were reached by the T2C team for results disclosure, initial medical and social needs assessment and provision of education (**Fig 2**); 74 (83%) reported Spanish as their preferred language. At the time of results disclosure and initial assessment, 27 (34%) had current symptoms compatible with COVID-19 (**Table 3**); 36 (45%) participants were without health insurance, including 4 participants who noted a recent lapse of health insurance coverage due to COVID-19-related job loss. Forty-four (55%) participants were without a primary care provider, while 36 (45%) had an established primary care clinic. There were 10 (12.5%) participants who said that they would be unable to safely isolate at their current address even with community support and home deliveries (either due to shared living spaces or because they were homeless) and they were provided a temporary room by the SFDPH at an isolation and quarantine hotel. The majority of participants (63%) stated that they would be able to isolate at home, but requested community-based support, including home deliveries; 20 (25%) participants said that they required no support to safely isolate at their current address (**Table 4**). Based upon initial assessment of the degree of support needed, 41 (51%), 34 (43%), 5 (6%) participants were classified as needing a low, medium and high amount of support, respectively.

**Table 4.**
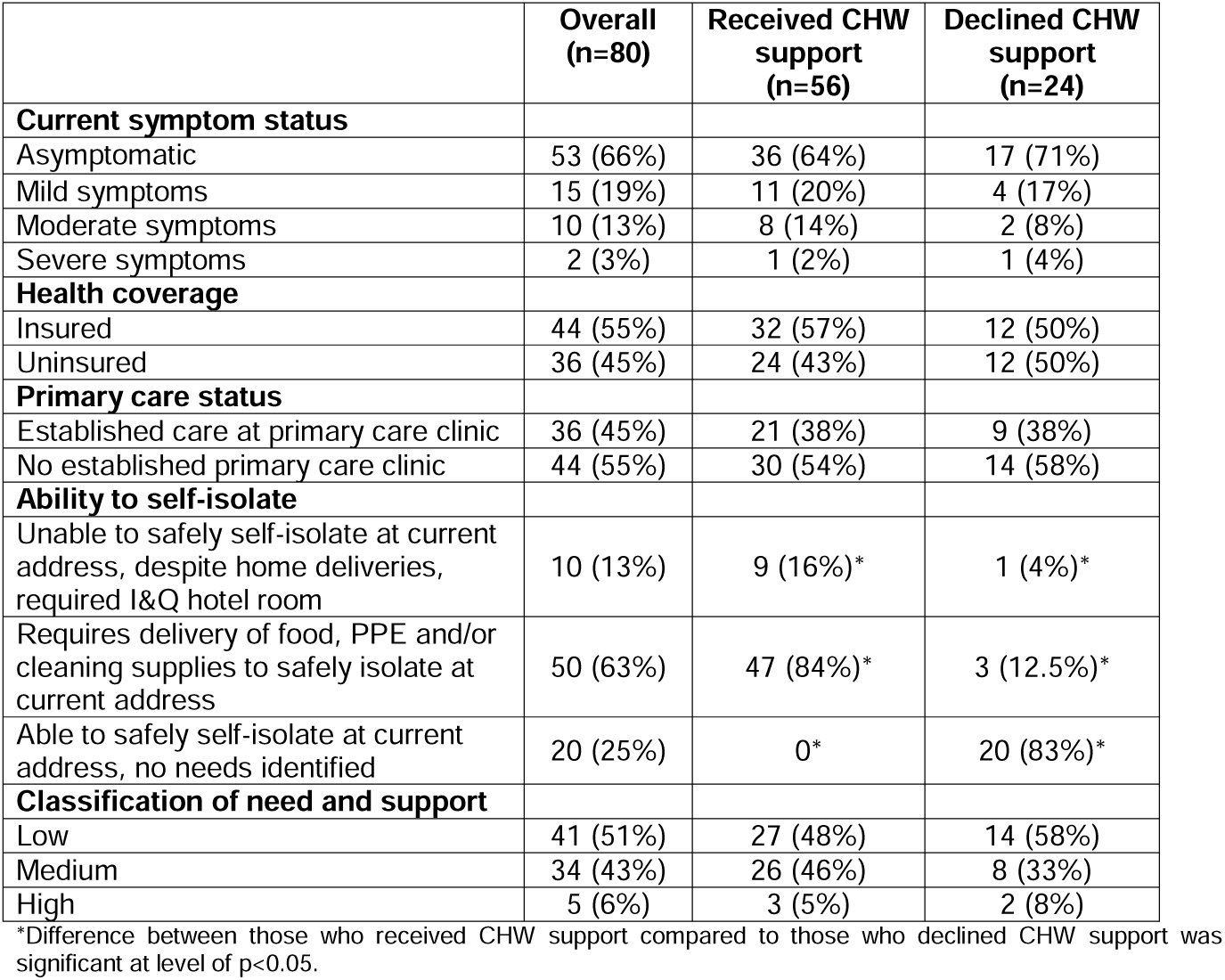
Characteristics and needs of COVID-19 positive participants identified during initial needs assessment.

**Fig 2.**
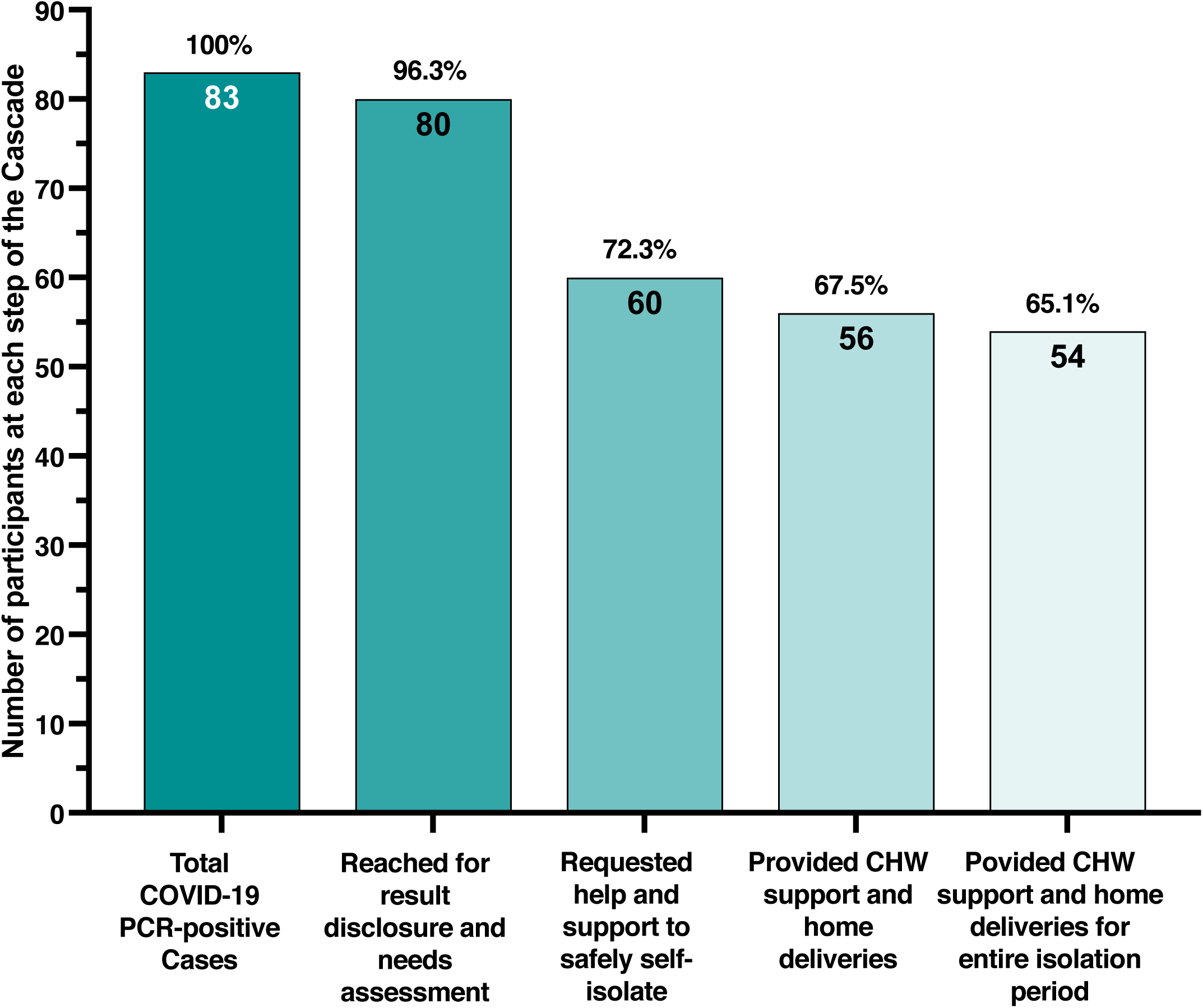
Cascade of enhanced community-based support among COVID-19 individuals

### Household contacts and contact investigation

All 80 COVID-19 positive patients reached for disclosure were asked details about additional household members, including name, age and whether they had been tested for COVID-19; COVID-19 positive individuals and their contacts were then connected with the SFDPH to facilitate routine case investigation and contact tracing. Participants were provided information on community sites where household members could go for free COVID-19 testing through the T2C team and also through standard SFDPH case and contact investigation services.

### Longitudinal support provided by T2C team

Each of the 80 participants who were reached for disclosure were regularly assessed by healthcare providers for new/progressive symptoms throughout the isolation period. Of 35 uninsured participants and 44 participants without primary care, 27 (77%) and 31 (70%), respectively asked for social work support to help link to care; 28 participants were reached by a T2C social worker and provided further information on health insurance enrolment and/or primary care establishment; for those who were interested, primary care appointments were directly scheduled on their behalf.

Of the 80 participants reached for disclosure and needs assessment, 60 participants requested support from CHWs to help safely self-isolate (**Table 4)**. CHWs were subsequently unable to reach three participants despite multiple attempts (median 6 attempts) and one participant no longer wished to be contacted by the T2C team. Therefore, 56 (67% of all COVID-19 positive participants) participants were reached by CHWs, provided social support and at least one home delivery (**Fig 2)**. Participants who received community-based support from CHWs were more likely to reside in densely populated households and live with other confirmed COVID-19 positive individuals compared to participants who declined community-based support (**Table 3)**; there was otherwise few differences observed among individuals supported by CHWs and those who were not (**Tables 3 and 4**).

## Implementation

### Acceptability

We found that the T2C model was highly acceptable to participants. Participants, especially those who received community-based, CHW support, expressed gratitude and a high level of satisfaction associated with the continued support offered beyond COVID-19 results disclosure. Participants reported that having Spanish speaking members of their community provide them support was very important to them and that they greatly appreciated the ongoing ability to ask questions and indicate new needs as they arose. Some participants directly commented on how reassuring it was to regularly communicate with and have access to healthcare providers, including social workers.

Participants came to know CHWs on a first name basis and over subsequent calls increasingly trusted them and shared information related to additional needs as well as household members not initially volunteered. This included 9 individuals who disclosed a larger number of household members than initially reported, 8 individuals who accepted supplies after initial reluctance and multiple refusals of material support (median 4 calls, range 3-5), and 6 persons who requested temporary relocation to a hotel room for isolation despite initially declining this service. The acceptability of the T2C model was high among both providers and CHWs.

The T2C Model was acceptable to providers and CHWs (**S1 Table**). Specifically, they reported that the T2C Model was an acceptable and appropriate way to address the many needs of low-income Latinx individuals with COVID-19, that it would be an appropriate model to address the needs of other low-income populations impacted by COVID-19 and that they would recommend the T2C Model to other providers and policy makers. Providers and CHWs also stated that they liked the approach and procedures used in the T2C model and that they greatly enjoyed working as a member of the T2C team (**S1 Table**).

### Feasibility

In general, the providers and CHWs felt that the T2C Model feasible to carry out as intended. Providers and CHWs became invested in the outcomes of the participants over the short time period of clinical follow-up. While significant time for initial and follow-up calls were sometimes required (at times up to 45 minutes), T2C providers and CHWs noted that the time investment became a strength of the model and allowed for the building of trust among T2C members and participants, allowing for a dynamic response to needs that might develop. They emphasized the importance of daily huddles and noted that when this did not occur, it created inefficiencies.

They reported that the T2C Model could likely be implemented in other settings – while not always easy to carry out, they felt strongly that it would be both possible and doable for other providers and CHWs to undertake (**S1 Table**) Providers and CHWs felt that providers and policy makers in other settings would be excited to implement a similar model in support of low-income individuals with COVID-19 and if implemented, it was a model that could be sustained over time (**S1 Table**).

### Fidelity

The majority of participants were called and reached either the same day (60/83, 72%) or within 24 hours (74/83, 89%) of COVID-19 positive result receipt to undertake disclosure, an initial needs assessment, and linkage to the SFDPH for contact investigation. T2C providers and CHWs provided longitudinal support to participants for the duration of the self-isolation period including ongoing education, symptom assessment, home deliveries as needed, and social support. During the demonstration project T2C CHWs made 250 daily phone calls (median 4 per participant, range 2-7) and 105 home-based deliveries (median 2 per participant, range 1-4) that included 300 bags of groceries.

### Effectiveness

Overall, participants communicated that regular check-ins and home deliveries of essential goods provided valued support during a very difficult period. Participants also noted that CHW support helped overcome feelings of loneliness and social isolation and provided them with increased confidence through ongoing education and reassurance.

Of the 36 (45%) participants without insurance at the time results were disclosed, 4 (11%) had documented health insurance one month later. Only 3 (7%) of 44 participants previously without a primary care provider established care at a clinic in the San Francisco Health Network within a month of follow-up. We identified three participants who developed severe symptoms of COVID-19 and were referred for urgent care or emergency room evaluation; one participant was hospitalized and none died. Notably, 54 (96%) of the 56 participants reached by CHWs were supported for the entire isolation period and received an exit package (**Table 2, Fig 2**). No participants articulated any stigma or discrimination experienced as a result of participation in the T2C model and providers and CHWs did not identify any unintended harms.

Initially, among 47 households, 246 total household members were reported. While few participants noted specific concerns about sharing information about all household members, through longitudinal CHW support, 9 of 47 households (19%) were found to have a higher number of household members than first reported. The total number of household members was found to be 284 - in one instance 20 additional household members were elicited than originally volunteered (30 total). Of 284 total household members, 118 (41.5%) were tested through the Unidos en Salud testing campaign. All remaining household contacts were referred by both the T2C team and SFDPH to free community COVID-19 testing.

## Discussion

We found that a novel, short-term care model designed to provide enhanced clinical and community-based support to socioeconomically vulnerable, COVID-19-positive Latinx persons following diagnosis and during isolation was feasible, acceptable, and reached a majority of individuals after a mass testing campaign. To our knowledge, the T2C model is the first model of its kind developed in partnership with community members and designed to address the specific needs during isolation and quarantine in a community disproportionately impacted by COVID-19. The care model augmented routine services provided by the SFDPH with additional provider-led and CHW-led components in order to holistically support the needs of low-income individuals during the required isolation period.

The majority of highly socioeconomically vulnerable COVID-19 participants requested direct support to safely self-isolate at their current address. This study suggests that the T2C model is both feasible and acceptable; if implemented, it could improve the ability of low-income COVID-19 positive persons to isolate and quarantine by directly addressing many of the barriers that they face. In order to halt COVID-19 community transmission and reduce its disproportionate impact among racial and ethnic minorities, “test, isolate and trace” strategies, must be coupled with a robust support component to facilitate early and effective isolation and quarantine among COVID-19 cases and their close contacts [7,8]. Without this promise of support to address the many barriers low-income, Latinx and other socioeconomically vulnerable persons face to safely self-isolate and quarantine, they are unlikely to undertake testing, even if provided low-barrier testing options, especially since COVID-19 can manifest with mild or no symptoms and the financial impact of isolation and quarantine is large. While most participants requested ongoing, community-based support, it is important to highlight that more than one quarter of low-income, Latinx COVID-19 positive participants declined enhanced CHW support. Participants who declined did not differ in any substantial way with regard to demographics, socioeconomic status, current symptomology or connectedness to primary health care than those who accepted. Further work is required to determine how to improve the uptake of the T2C Model in order to maximize its reach among all socioeconomically vulnerable persons who are likely to benefit from its support.

There are a number of lessons that we learned during the course of the demonstration project, several of which could potentially be applied to improve the efficiency and effectiveness of the T2C model. First and foremost, the longitudinal follow-up provided by Spanish-speaking CHWs and care providers to participants was a key feature of the T2C model’s design that allowed us to develop trust with participants over the course of two weeks through repeated phone calls and/or texts. Many participants were initially reluctant to accept help and services; additionally, some participants did not initially anticipate the challenge of isolating for 10 days and declined services, including isolation hotel rooms, on that basis. Due to relationships formed with CHWs combined with ongoing assessments, many participants ultimately accepted social and material support and two-thirds of all COVID-19 positive participants were provided ongoing support from CHW, including home deliveries, for the entire duration of self-isolation. We therefore would expect that vulnerable persons in other settings might initially decline help and services, but would strongly benefit from and may eventually accept support after multiple offerings. Trust of CHWs also allowed greater information about the household to be elicited and CHWs often learned that there were more household members than initially volunteered. This allowed us to more optimally support the true needs of the entire household.

Team members and participants strongly highlighted the need for improved communication and integration of T2C teams within the DPH. Due to the initial T2C model design, participants sometimes received calls from both the T2C team and SFDPH (for contact tracing) in the same day, which at times caused confusion and frustration among participants as they were not always clear who was calling, how one caller was from a different organization from the prior caller, and the purpose of each call. Full integration of community-based organizations and CHWs into the DPHs’ case investigation and contact tracing services, with the addition of the longitudinal services of the T2C model over the duration of isolation, would enhance communication and optimize trust and support for community members [25,38].

Nearly half of COVID-19 positive participants were without health insurance and almost two-thirds did not have an established primary care provider. While we sought to provide dedicated support to facilitate establishment of health insurance and linkage to primary care, we found that after one month only 11% previously uninsured participants were now insured and 7% of those previously without a primary care provider were newly established in care. Notably, 23% of uninsured participants and 30% of those without primary care declined social work support to link to care. While care linkage estimates may be underestimated due to our inability to ascertain who established care outside of the main public health network, possible delays in scheduling related to COVID-19 and the relatively short follow-up period (one months), this still suggests that an important proportion of participants did not establish primary care. Among COVID-19 positive participants who tended to be younger (72% 18-50 years old), and were likely to be asymptomatic (66%), competing priorities may have made them less likely to invest the time required to establish care due to a lack of perception of ongoing health needs. Language concordance and immigration concerns may have also been important barriers. At the time of the study, the new patient hotline for the main public health network’s recorded greeting was only in English, but is now also in Spanish and Cantonese. Additionally, we conveyed to participants that seeking care does not count as a public charge, but this may still have been a barrier to establishing care; under current U.S. law, individuals who have received certain public benefits are deemed likely to become primarily dependent on the government for their basic needs are known as a “public charge” and are not eligible to become a legal permanent resident or receive a temporary visa [39]. Interventions to improve linkage to primary care after a COVID-19 diagnosis warrant further assessment. Possible interventions for further study include: enrollment into primary care at the time of testing, enhanced patient navigation such as having CHWs join participants’ calls to the new-patient enrollment phone number, direct outreach from the local clinic or health network, or reserving new appointment slots in local clinics to facilitate rapid intake and care engagement.

We identified challenges to reaching household members and other close contacts of Latinx individuals to undertake contact investigation and COVID-19 testing due to fear and distrust, but found that some of these barriers can potentially be overcome by longitudinal support and relationship building. Due to the high transmissibility of SARS-COV-2 and the close and prolonged contact among household members, such individuals are at extremely high risk of COVID-19 and should be prioritized for contact investigation – as evidenced by our finding that 61% of positive participants shared a household with at least one other confirmed case. Through the establishment of trust, the CHW-led T2C team found that nearly 20% of households had more household members than first reported. This suggests fear and/or reluctance among some Latinx individuals to report all close contacts to public health authorities, possibly due to immigration concerns (documentation status, Public Charge Rule) and concerns about impact on employment status, concerns which may undermine case investigation and contact investigation efforts. Furthermore, although all participants and their close contacts were provided information regarding free COVID-19 testing sites, further data is needed to assess the uptake of testing among household contacts. Moreover, interventions to expand low-barrier, community-based COVID-19 testing including self-test kits, mobile testing, or providing direct support to access existing services [6,40] coupled with surveillance systems to monitor and ensure support testing uptake among all household and other close non-household contacts, are needed.

Our results have several important implications for future COVID-19 response efforts. As demonstrated here, the multicomponent T2C model may offer an effective approach that can be implemented in other settings to support socioeconomically vulnerable individuals during isolation who are disproportionally impacted by COVID-19. While the T2C model can and should be adapted to the needs of a local community including other vulnerable populations, we believe that the CHW-led, community-based component - offering social support, ongoing assessments and home-deliveries - are major strengths of the T2C model and therefore represent core components. This concords with several prior studies that have shown CHW to be associated with positive health outcomes for a number of health conditions [9,11,12,17].

Despite multiple levels of support, many individuals still expressed an extremely strong need for financial support to assist with rent and cellphone bill payments as well as other expenses during isolation. Our study directly contributed to policy change in the city, with the establishment of the ‘Right to Recover Program’, which provides eligible individuals in San Francisco with COVID-19 with two weeks of wage replacement to allow them and their families to safely isolate and quarantine [41]; notably, the program does not require a formal application and does not ask about one’s citizenship or immigration status. We believe that low-barrier financial assistance, along with this comprehensive care model, should be an essential commitment made to support all vulnerable COVID-19 positive community members during the isolation period through the pandemic. Such programs are likely to increase the willingness of communities to undergo testing if they know they have access to funds to offset their lost wages.

Our study has some limitations. First, assessment of implementation outcomes among COVID-19-positive participants was limited to informal interviews; nonetheless, these still provided important information related to the acceptability of the T2C and will inform formal qualitative research as part of future evaluations of the T2C Model. Additionally, we were unable to directly assess adherence to isolation and quarantine among participants and their households; therefore, we could not directly assess whether the T2C Model was effective in enabling individuals to more closely adhere to recommended public health guidance. Reliable and validated approaches for monitoring adherence to (and appropriate support for) isolation and quarantine among vulnerable populations for whom mobile device tracking may not be acceptable are currently lacking and represent an important public health priority to optimize the effectiveness of COVID-19 test, isolate and trace strategies [8]. Finally, the T2C model was designed to address the specific needs of low-income Latinx persons with COVID-19 in the Mission District of San Francisco, California and thus our findings may not be generalizable; however, all T2C providers and CHWs felt that the T2C Model could be implemented in other settings and could be adapted to better support the needs of other low-income individuals.

In conclusion, the T2C model to support low-income individuals after a COVID-19 diagnosis was found to be highly acceptable to participants, feasible to undertake and, through direct and ongoing multilevel support, effective in supporting low-income Latinx individuals and their households through the period of self-isolation and quarantine. To further improve the effectiveness of this model, improved integration with public health services coupled with expansion of tailored, low-barrier COVID-19 testing options for close contacts is recommended.

## Data Availability

The data will be held in a public repository and will be made available after acceptance of the manuscript.

## Acknowledgements

We thank the community members from the census tract studied for their generous participation in our study during San Francisco’s shelter-in-place ordinance. We thank all members of the San Francisco Latino Task Force for COVID-19 who led who helped design the Test-to-Care Model and supported community outreach and education. We thank Gaynor Siataga for helping with home deliveries of essential goods. We gratefully acknowledge the contributions of Zuckerberg San Francisco General Hospital Ward 86 providers and staff who provided support for the Test-to-Care Model, including Jon Oskarrson, Mary Shiels, Erin Pederson, Myriam Beltran, Sandra Torres, Fatima Ticas, Tamareh Abualhsan, Helen Lin, John Szumowski, and Francesco Sergi. We also thank Stacie Powers for her generous donation of the Brava Theater space to operate as headquarters for the community-based Test-to-Care Team. We also thank Grocery Outlet for their generous donation of groceries to support this demonstration project.

## Funding Statement

The study was supported by the Chan Zuckerberg Biohub, UCSF, and a Program for Breakthrough Biomedical Research award. The funders had no role in study design, data collection and analysis, decision to publish, or preparation of the manuscript.

## Ethics Statement

The UCSF Committee on Human Research determined that the study met criteria for public health surveillance, program evaluation and quality improvement activities and was exempt from IRB oversight.

## Notes

### Competing Interest Statement

The authors have declared no competing interest.

